# FHIR-DHP: A Standardized Clinical Data Harmonisation Pipeline for scalable AI application deployment

**DOI:** 10.1101/2022.11.07.22281564

**Authors:** Elena Williams, Manuel Kienast, Evelyn Medawar, Janis Reinelt, Alberto Merola, Sophie Anne Ines Klopfenstein, Anne Rike Flint, Patrick Heeren, Akira-Sebastian Poncette, Felix Balzer, Julian Beimes, Paul von Bünau, Jonas Chromik, Bert Arnrich, Nico Scherf, Sebastian Niehaus

## Abstract

**Background:** Increasing digitalisation in the medical domain gives rise to large amounts of healthcare data which has the potential to expand clinical knowledge and transform patient care if leveraged through artificial intelligence (AI). Yet, big data and AI oftentimes cannot unlock their full potential at scale, owing to non-standardised data formats, lack of technical and semantic data interoperability, and limited cooperation between stakeholders in the healthcare system. Despite the existence of standardised data formats for the medical domain, such as Fast Healthcare Interoperability Resources (FHIR), their prevalence and usability for AI remains limited.

**Objective:** We developed a data harmonisation pipeline (DHP) for clinical data sets relying on the common FHIR data standard.

**Methods:** We validated the performance and usability of our FHIR-DHP with data from the MIMIC IV database including > 40,000 patients admitted to an intensive care unit.

**Results:** We present the FHIR-DHP workflow in respect of transformation of “raw” hospital records into a harmonised, AI-friendly data representation. The pipeline consists of five key preprocessing steps: querying of data from hospital database, FHIR mapping, syntactic validation, transfer of harmonised data into the patient-model database and export of data in an AI-friendly format for further medical applications. A detailed example of FHIR-DHP execution was presented for clinical diagnoses records.

**Conclusions:** Our approach enables scalable and needs-driven data modelling of large and heterogenous clinical data sets. The FHIR-DHP is a pivotal step towards increasing cooperation, interoperability and quality of patient care in the clinical routine and for medical research.

## Introduction

The increasing digitalisation of healthcare creates vast amounts of clinical data that are collected and stored in an Electronic Health Record (EHR). Patient information from all medical domains is captured in diverse sets of data recorded in standalone systems. With the prevalent use of EHRs in healthcare organisations, there is abundant opportunity for additional application of EHR data in clinical and translational research. For instance, such data can be used to develop artificial intelligence (AI) algorithms which have the potential to transform patient care and medical research. Resource intensive and inefficient clinical workflows could be optimised by the analysis of historical data with AI applications (1,2). In particular, the time-consuming and high-priced process of identifying and enrolling the right patients into a clinical trial manually can be reduced significantly by automation (3,4). However, the exchange of medical data remains limited due to the lack of data interoperability between healthcare providers, owing to outdated IT infrastructure, inconsistencies in data formats, poor data quality, inadequate data exchange solutions and data silos (5,6). To achieve data interoperability, the following steps must be incorporated: i) integration of isolated data silos, ii) safe exchange of data and iii) effective use of the available data (7). Each of these operations includes database schema matching (8) and schema mapping (9), which allow translation of the relationships between the source database and the target data standard.

Employing a harmonised data format will facilitate the exchange of medical data, enabling wide-ranging data-driven collaborations within the private and public healthcare sectors. Data interoperability requires EHR data to be structured in a common format and in standardised terminologies. Standardisation is often performed by adopting the Health Level 7 (HL7) Fast Healthcare Interoperability Resources (FHIR) model (10), which is supported by numerous healthcare institutions and vendors of clinical information systems (11). FHIR is an international industry standard with the benefit of integrating diverse sets of data in well-defined exchangeable segments of information, which are known as FHIR resources. Therefore, FHIR facilitates interoperability between healthcare organisations and allows third-party developers to provide medical applications which can be easily integrated into existing systems. FHIR enables the harmonization of data and thus allows standardized data processing and also the rollout of AI applications across different clinics and hospitals regardless of which information system they use. Therefore, FHIR forms an important component for the scalable development and deployment of AI in clinics and hospitals.

However, to apply AI, the input data needs to be adapted to the AI algorithms. The conventional AI frameworks such as Tensorflow (14) and Pytorch (15) require data to take a tensor form which is a vector or matrix of n-dimensions that represents various types of data (e.g., tabular, time series, image, text). FHIR facilitates the application of AI in medical domain as it provides needed interoperability for a standardised access of EHR data. FHIR format’s multi-layered nested structure requires case-specific data pre-processing to use it for AI algorithms. Depending on the AI application and the chosen data source, a custom data preprocessing pipeline needs to be designed, which leads to diminished AI scalability. Up to the present time, a number of studies have attempted to solve this problem. Prior research addressed this problem in different forms, but focuses on individual use cases and thus constrains the basic idea of FHIR to be independent of the use case.. There have been a few attempts to flatten the hierarchical FHIR structure and transform it into NDJSON-based data format (16) or tabular format saved in CSV files (17). Such formats are more AI-friendly as they represent the data in a more accessible and standardised form for an application of common AI frameworks. Nonetheless, the NDJSON-based FHIR data transformation approach (16) does not provide data selection criteria and filtering capabilities. The approach presented in (17) requires expert knowledge of *FHIRPath* query language.

In this paper, we address the challenge of data interoperability in the healthcare sector by proposing a FHIR Data Harmonisation Pipeline (DHP) that provides EHR data in an AI-friendly format. The newly developed FHIR-DHP represents a data workflow solution that includes the aforementioned operations such as data exchange, mapping, and export. Data privacy is a delicate topic in healthcare and is of great ethical concern (18). Given the degree of automation, such pipeline should allow preprocessing of unseen data in an isolated hospital environment, which makes the harmonisation privacy-preserving. In this setting, direct access to the sensitive data would not be required to run the standardisation pipeline. FHIR-based data preprocessing pipelines have already been implemented in different contexts: as electronic data capture (12), as a natural language processing tool (13) and as a standardisation protocol based on the Resource Description Framework (RDF) (6). Despite their immense benefit of processing EHR data, existing approaches are limited to specific use cases or require considerable data preparation to perform standardisation. Moreover, their final output is not easily accessible by common data preprocessing tools and thus hinders the application of AI.

## Methods

### FHIR-DHP Development

In our work, we propose a generic solution to harmonise hospital EHR data. The FHIR-DHP was designed based on the Extract-Transform-Load (ETL) framework (19) in which the data is pulled out (i.e. queried) from diverse sources, processed into the desired format and loaded into a data warehouse, namely the “patient-model DB”. As the hospital database (DB) contains highly sensitive patient data, it is located behind the hospital’s security infrastructure and is completely isolated from outside access. Therefore, an edge-computation solution was designed, bringing the FHIR-DHP into the hospital’s own infrastructure. The edge-computation solution represents a set of frameworks which perform data querying, preprocessing, storage and export. In this setting, direct access to the sensitive data is not required to run the standardisation pipeline. The queries to the data are defined beforehand based on the database documentation.

To bring the data into a harmonised form we used Fast Healthcare Interoperability Resources (FHIR) data model which is applied by mapping the relationships between the source database and the desired data standard. The FHIR standard is straightforward to implement because it provides a choice of JavaScript Object Notation (JSON), Extensible Markup Language (XML), or Resource Description Format (RDF) for data representation. The mapping pipeline was developed in Python programming language to translate queried hospital data into matchig FHIR concepts and save the resulting resources in JSON format. The conversion to FHIR was designed to only support a core standard of the FHIR format to allow generic data preprocessing.

Syntactic validation of FHIR resources is necessary in the remote data standardisation scenario to prevent errors. For instance, conversion of data types can sometimes lead to wrong values, especially with date features. Automatic syntactic validation allows logging of occurred errors and improvement of standardisation pipeline when working with unseen data. After the mapped data is validated, FHIR resources should be sent to the database for storage to allow fast and easy retrival of preprocessed data for AI applications.

In the final stage of data export, we designed the output that provides the benefits of the original FHIR format with a high level of clinical detail, yet which is also easily accessible for computational tools. Moreover, we wanted to restructure the data representation in a way which supports efforless data selection and filtering capabilities and which would not require knowledge of *FHIRPath* query language. Consequently, such output format would enable smooth conversion of data into a “tensor” format required by conventional AI frameworks.

### FHIR-DHP Validation

To demonstrate and evaluate how the FHIR-DHP works, we used the openly available Medical Information Mart for Intensive Care IV (MIMIC IV) database (20). MIMIC IV includes patient data from over 40,000 individuals admitted to intensive care units at a tertiary academic medical center in Boston, MA. We selected a wide range of tables from MIMIC IV which cover most of the events occurring during the hospital stay as well as core patient details, information about admissions and hospital transfers (further referred as core tables). The event tables include laboratory results, diagnoses, prescriptions and other details as shown in **Table 1**. MIMIC IV includes so-called reference tables containing matching dictionaries with medical terms which are used in the hospital records.

**Table 1.**
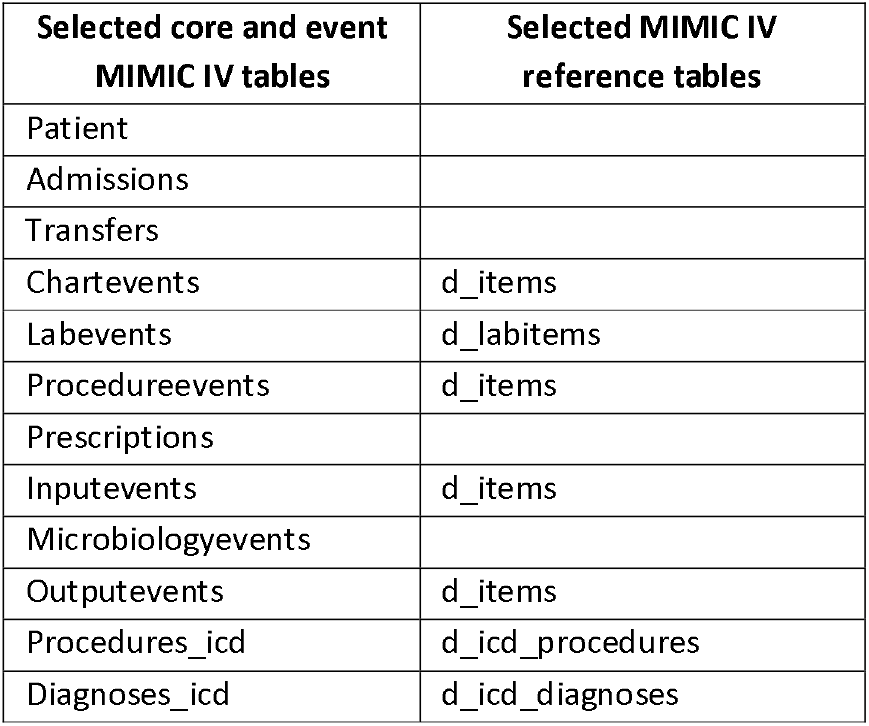
The table lists selected core and event MIMIC IV tables as well as the reference dictionary tables which were merged together with core and event tables for FHIR mapping.

The selected tables were mapped to FHIR standard. Automatic semantic validation is unfeasible, so two of the authors manually validated the mapping semantics independently of each other. There are many tools which perform automatic syntactic validation, such as the Python-based package fhir.resources used herein (21). To evaluate the exporting of data from the patient-model DB, we retrieved diagnoses records.

## Results

### FHIR-DHP Architecture

The approach presented herein represents a scalable protocol for harmonising hospital EHR datasets based on five stages from data query to data export in a standardised format.

#### 1. Querying data from the hospital database

To connect the FHIR-DHP pipeline to the hospital DB, a communication server is employed. This server runs all necessary queries to retrieve the patient data. The query execution can be run at regular intervals as well as in batches of patients, so as not to overload the data pipeline. Furthermore, the queries pre-structure the data according to their semantic relations before proceeding to data mapping.

#### 2. Mapping data to FHIR

FHIR allows describing data formats and elements which are recorded as “resources” and an application of a programming interface (API) for exchanging EHRs. To perform the mappings, semantics of features from the source database and FHIR concepts are explored as well as relationships between the data tables. Consequently, the mappings between the database tables and FHIR resources are defined. Features where a matching FHIR concept is not found are excluded. The resulting FHIR resources are then saved in JSON format.

#### 3. Syntactic validation of FHIR mappings

During validation, mapped data is ensured to have the correct data types as well as the syntactic format where the hierarchy is maintained and entries follow FHIR standard specifications. All mappings are validated first during the development stage to identify structural errors and data type inconsistencies. A validation algorithm is incrorporated into the pipeline to confirm the correctness of transformed data in the remote data standardisation scenario.

#### 4. Transferring FHIR resources to patient-model DB

The database of choice for the patient-model is Postgres (22) which is an open-source relational database management system (RDBMS) featuring SQL compliance and storage of JSON documents. PostrgreSQL allows handling both small and large workloads. The database for FHIR resources is used to harmonise the locally available data only once to allow further application of various medical AI-based solutions. The data is stored according to FHIR resource type where each resource is saved in a separate JSON structure.

#### 5. Exporting data into custom JSON format

To export the data from the patient-model DB, the selection is performed by outlining the tables and features of interest in a configuration file which is then used to determine which data is queried from the patient-model DB. Following that, the data is exported into the custom JSON file adhering to specific formatting rules in respect of its key-value structure. To create a custom JSON structure, *FHIRPath* queries were written to retrieve all elements from FHIR resources. Such transformation flattens the hierarchical structure of FHIR resources and makes the data more accessible for common data preprocessing tools. The final flattened output does not require expert knowledge of *FHIRPath* query language and supports effortless data selection and filtering. The resulting file allows uncomplicated conversion of data into a “tensor” format required by conventional AI frameworks and fast data selection based on four keys: feature_name, table_name, value and metadata.

In **Figure 1**, we demonstrate how the FHIR-DHP recodes nested FHIR syntax to more accessible features in an AI-friendly format. Example FHIR concepts from an Observation resource are given in **Figure 1a** where the code’s entity “text” defines the record or measurement label. The entity “text” is often duplicated in the item “display”. However, depending on the coding system this “display” item can change, whereas “text” always stays the same and is therefore used as a feature name. The information from the FHIR resource is grouped into four concept-keys such as feature name (ex. “Blood pressure”), value (ex. “114”), table name (ex. “observation”) and metadata (**Figure 1b**). For a given FHIR resource type, the metadata may include concepts such as dates, references, coding system details, resource ID amongst other things. As an output, feature names together with a corresponding value and available metadata are provided in a custom JSON structure (**Figure 1c**). The defined format allows uncomplicated data selection and aggregation based on resource type (ex. “table_name”), feature name and value. Additional information in a standardised format can be easily accessed from the metadata key and allows further data manipulation.

**Figure 1.**
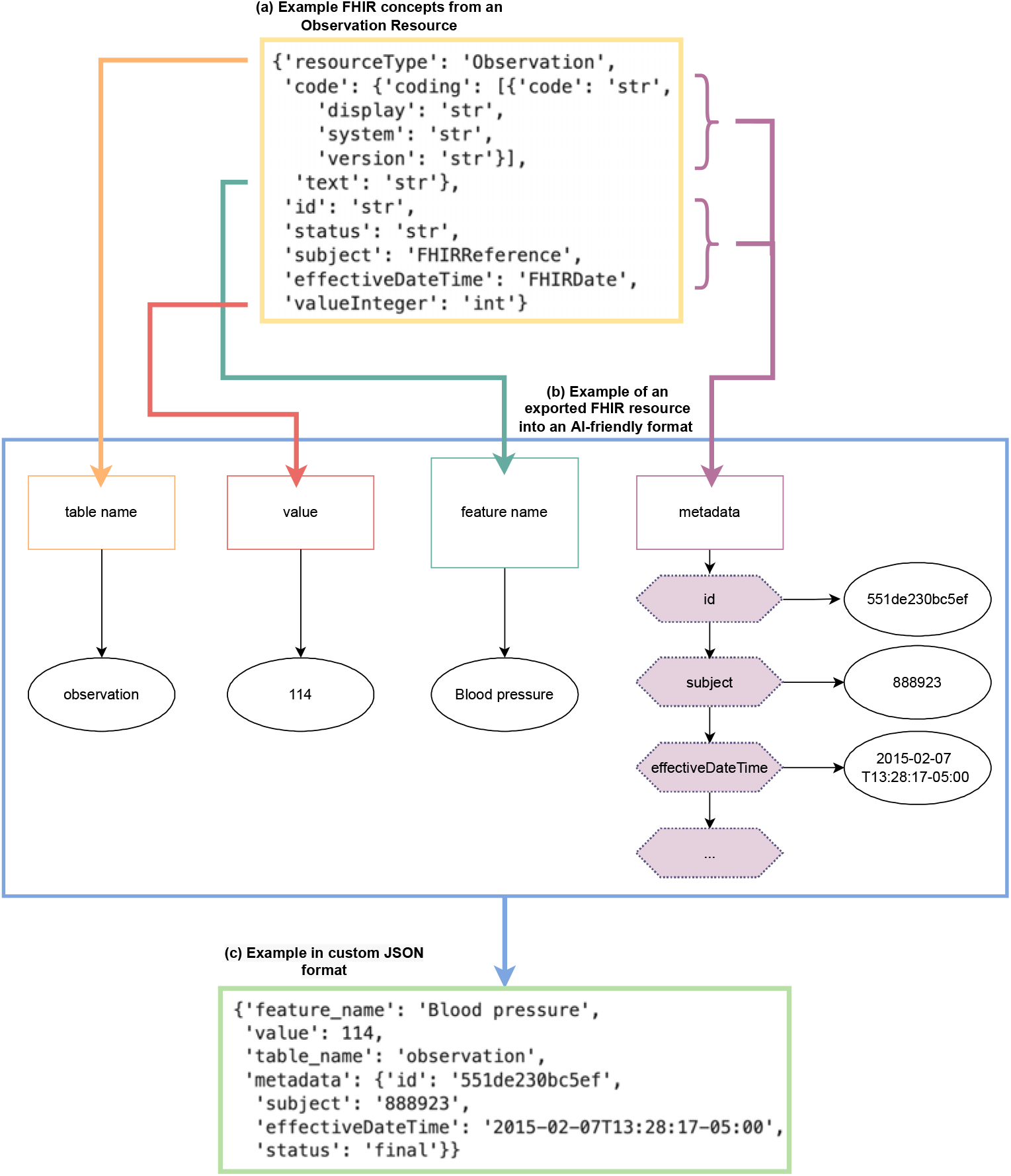
Conceptual overview for an exemplary FHIR structure and hospital record which are transformed from FHIR standard to an AI-friendly format.

### FHIR-DHP Validation

The MIMIC IV data was queried accordingly to the defined FHIR mappings. The core and event MIMIC IV tables were merged with reference tables to contain complete description of the hospital records. As a result, the data was grouped and restructured into the information blocks required in FHIR standard. Manual independent validation of the mapping semantics resulted in slight descrepancies which were subsequently resolved to adhere closely to the FHIR standard. The automatic syntactic validation allowed prompt verification of standardisation operations.

**Table 2** shows to which FHIR resources the MIMIC IV tables were mapped. The largest proportion of tables (4 out of 12 tables) were mapped to the *Observation* FHIR resource type which included lab, microbiology, output and charted events collected throughout the patient stay. The information on admissions and transfers was translated into the *Encounter* FHIR resource (2 out of 12 tables). Procedure events and ICD codes (2 out of 12 tables) were stored in the *Procedure* FHIR resource. Given that the prescriptions table contains medication requests (1 out of 12 tables) and inputevents table holds records of medication administration (1 out of 12 tables), these tables were mapped to corresponding FHIR resource types. Finally, the *Condition* FHIR resource was used to map the table with patients’ diagnoses details (1 out of 12 tables).

**Table 2.** Overview of mappings performed on the selected MIMIC DB tables to FHIR resource types.

| MIMIC IV DB | FHIR Resource Type |
| --- | --- |
| Patients | Patient |
| Admissions | Encounter |
| Transfers | Encounter |
| Chartevents | Observation |
| Labevents | Observation |
| Procedureevents | Procedure |
| Prescriptions | MedicationRequest |
| Inputevents | MedicationAdministration |
| Microbiologyevents | Observation |
| Outputevents | Observation |
| Procedure_icd | Procedure |
| Diagnoses_icd | Condition |

In **Table 3**, we demonstrate how the mapping of the MIMIC IV “diagnoses_icd” table to *Condition* FHIR resource was conducted. Multiple columns of the “diagnoses_icd” table such as “icd_code”, “icd_version” and “long_title” were mapped to FHIR “condition.code” concept, which has a nested structure and provides keys to store the exact ICD code, version of the coding system and the code title. The full diagnosis title was mapped both to the “display” and “text” entities.

**Table 3.**
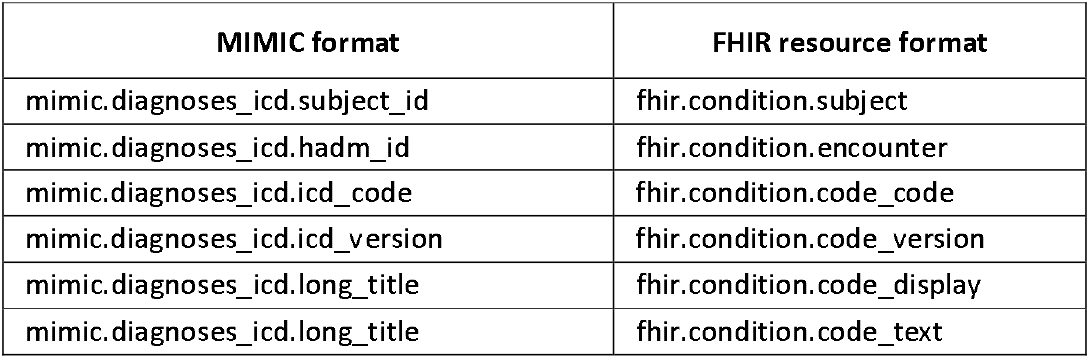
Mapping of “diagnoses_icd” table to Condition FHIR resource.

| MIMIC format | FHIR resource format |
| --- | --- |
| mimic.diagnoses_icd.subject_id | fhir.condition.subject |
| mimic.diagnoses_icd.hadm_id | fhir.condition.encounter |
| mimic.diagnoses_icd.icd_code | fhir.condition.code_code |
| mimic.diagnoses_icd.icd_version | fhir.condition.code_version |
| mimic.diagnoses_icd.long_title | fhir.condition.code_display |
| mimic.diagnoses_icd.long_title | fhir.condition.code_text |

**Figure 2** shows an example of how queried diagnoses records are harmonised to an AI-friendly format. The standardisation follows the described FHIR-DHP stages. At first, the raw data from tables “diagnoses_icd” and “d_icd_diagnoses” is queried (**Figure 2a**) and merged accordingly to the defined FHIR mappings. Then the features are renamed as defined in **Table 3** for FHIR Condition resource and required entities such as “resourceType” and “id” are created (**Figure 2b**). Finally, the values are placed into a nested FHIR structure (**Figure 2c**), and subsequently the data is transformed into JSON format (Figure 2d), which can be automatically validated (**Figure 2e**) and saved in the patient-model DB. When the resource is not approved in terms of its syntactic quality, e.g. data type, nested structure or cardinality, an error is raised which prevents further saving of this resource in the patient-model DB (**Figure 2e**). Otherwise, the resource is transferred into a storage (**Figure 2f**) and the requested data is exported in a custom AI-friendly JSON format (**Figure 2g**).

**Figure 2.**
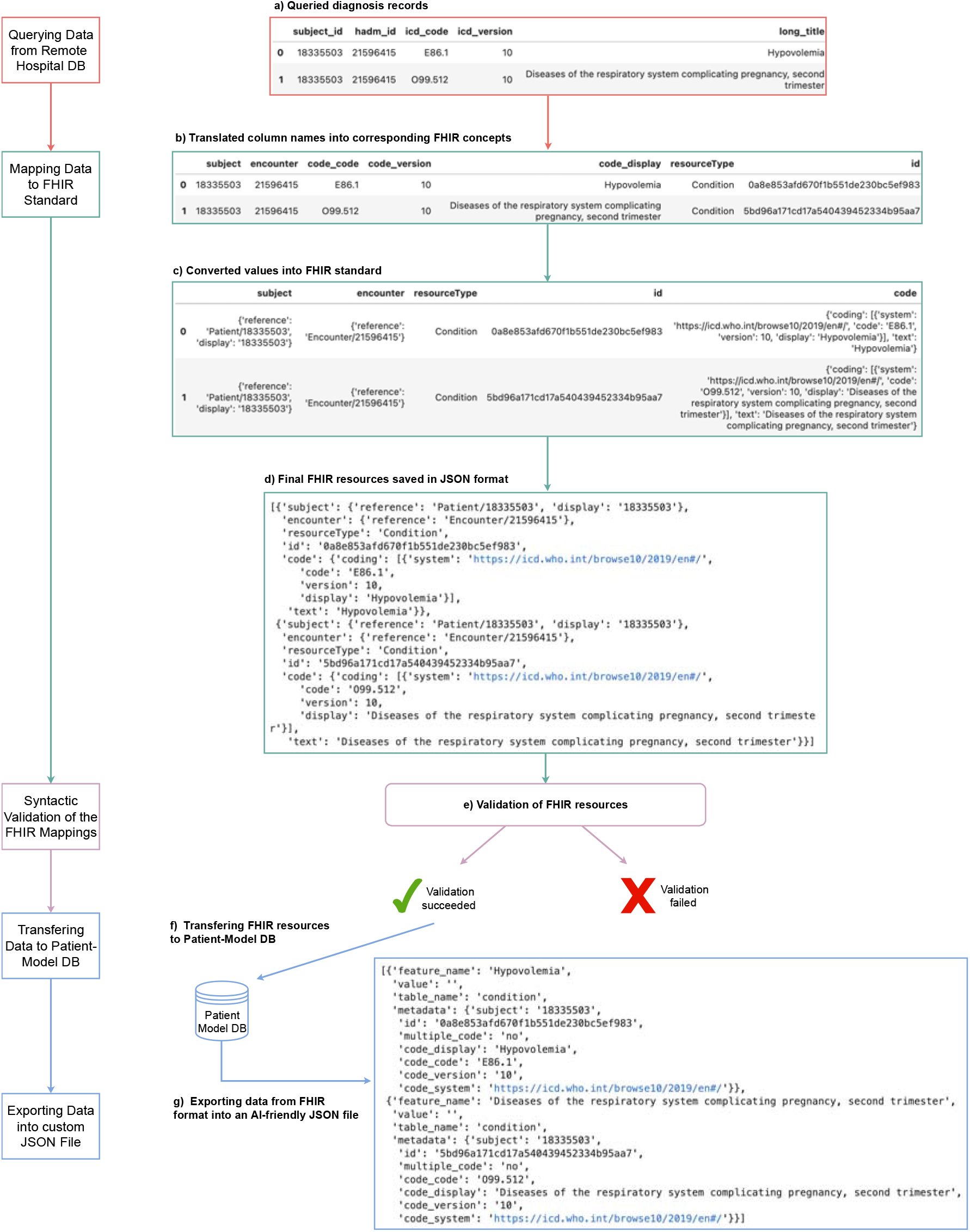
Flow chart showing an example diagnoses data being processed through the five stages in FHIR-DHP. The first stage includes querying of the diagnoses records (a), at the second stage the data is mapped to FHIR standard (b-c), and the third stage carries out the syntactic resource validation. If the FHIR resource is successfully validated, it is being transferred into the patient-model DB (f) and then exported in a custom AI-friendly JSON format (g).

We provide an example of further two-step transformation of harmonised example diagnoses data to a “tensor” format in Supplementary Material, chapter A.

## Discussion

Harmonisation of EHR data is a crucial step towards increasing cooperation, interoperability and quality of patient care in the clinical routine and medical research. To drive harmonisation of medical data forward, we developed the FHIR-DHP and evaluated it on key MIMIC IV tables. A detailed example of data standardisation was presented for clinical diagnoses records from the MIMIC IV database. The FHIR-DHP allows querying of health data in an isolated environment employing an edge-computation solution and a communication server which retrieve patient data and pre-structure it for further mapping to the FHIR standard. A validation step ensures syntactic compliance and initiates transfer of formatted data to the patient-model DB. The data export provides FHIR resources in a custom JSON file format.

Owing to the FHIR format’s multi-layered nested structure, its accessibility for AI algorithms is low as it requires transformation into a format compatible with common data preprocessing tools. Up to the present time, a number of studies have attempted to solve this problem. However, the final output of these studies has not supported data selection criteria and filtering capabilities (16) and requires expert knowledge of *FHIRPath* query language (17). Here, we introduce a custom JSON format which represents a higher level of abstraction to support easier data selection based on four keys: feature_name, table_name, value and metadata. Moreover, the newly developed JSON structure fits the expected data format of common data preprocessing frameworks, which are designed to work efficiently with tabular data. As a result, the ouput presented facilitates generic and fast deployment of AI and patient cohort identification algorithms.

In comparison to (12,13), the details of FHIR-DHP execution inside the hospital environment to protect data privacy are discussed. This step, though crucial, is often omitted and left out of the published standardisation protocols. The edge-computation solution sets up the FHIR-DHP in a privacy-preserving way where preprocessing of the patient-related data is performed inside the hospital and is completely isolated from outside access. So-called federated learning (FL) framework (23) can be integrated into FHIR-DHP workflow to run algorithms locally using the data on the on-premise component in the respective hospitals and to merge model parameters centrally in the cloud without any patient data leaving the hospital. The FL framework requires data to be in a consistent format across various hospital systems. The developed pipeline achieves such a format and enables scaling of AI applications. Furthermover, given the degree of automation, the setup of the pipeline facilitates preprocessing of unseen data in an isolated hospital environment, which makes the harmonisation privacy-preserving.

To the date of publication of this paper, there are only two studies attempting to perform mapping of MIMIC IV database (24,25). In (24), the mapping was performed on fewer tables than our approach (8 versus 12 tables). The FHIR mappings from (25) have been recently released and were not yet widely validated. Similarly to (12,13,24), FHIR-DHP includes verification of the performed FHIR mapping which is essential to ensure validity of data transformation. An automated syntactic verification of translated to FHIR data is crucial to adhere to FHIR version updates. Moreover, in comparison to (12,13,24), FHIR-DHP represents a generic approach to standardise EHR data and can be applied to various hospital database systems.

The FHIR-DHP allows integration into the hospital data management system which facilitates the development and application of advanced AI and patient cohort identification algorithms without compromising on data privacy protection laws. With the introduction of the FHIR-DHP into the hospital environment, a number of patient stay parameters can be potentially optimised using AI-based algorithms. For example, the length of stay as well as mortality could be reduced (26) and patients suitable for trial treatment could be automatically and efficiently identified (27). In consequence the financial impact on medical providers in respect of personnel time and resources would decrease considerably. The FHIR-DHP aims to bring healthcare closer to digital transformation and thus towards Healthcare 4.0 (28) by making EHR data usable “from bedside-to-bench”. By inverting the idea of translational research, in contrast to “from bench-to-bedside”, accessing the full potential of medical big data with AI will further inform and advance basic research.

There are several limitations that we would like to emphasise. FHIR-DHP only works with a core standard of the FHIR format. Those core FHIR resource types have a bounded set of concepts which presents a constraint to mapping accuracy. Although the standard resources can be expanded using profiling technique or FHIR extensions, the use of those would make the FHIR-DHP less generic. Hence, we implemented the mapping using only the standard FHIR resources and omitted some of the MIMIC IV data features which did not have a matching concept in FHIR. Additionally, the FHIR mapping step is subject to the extent of the detail of the database documentation used to infer semantic and syntactic properties of the data. A solution for an automatic concept recognition can potentially solve this problem. The existing approach in (6) is limited to a small number of FHIR resources and requires an extensive data preparation. Further experiments in this direction could alleviate the concept matching problem and the requirement for a detailed database description. Moreover, the validation and robustness of FHIR-DHP needs to be tested on other EHR datasets to evaluate its generic setup. In addition, to validate the FHIR-DHP compatibility with machine learning pipelines, further experiments are needed.

The proposed FHIR-DHP pipeline highlights the therein featured essential data standardisation stages and holds the potential to becoming an interoperable harmonisation system with an AI-friendly data format. FHIR-DHP enables interoperability and cooperation between clinical institutions, rapid patient cohort identification for clinical trials and unlocks the potential of big medical data.

## Conclusions

We provide a comprehensive approach to transforming unstandardised EHR data into a harmonised multi-layered nested FHIR format and then to a more readable, more efficient AI-friendly JSON structure. We developed a five-stage data harmonisation pipeline, which includes validation checks. The AI-friendly format of patient data allows generic and fast integration of both AI and patient cohort identification algorithms. Harmonised and standardised health care data is of great value to advancing efficiency in big data processing, cooperation and multi-center data exchange in the clinical sector, in order to boost medical research, patient care and clinical trial cohort identification. The next steps would include validating our approach in a hospital environment and applying privacy-preserving FL framework to make use of advanced AI deployment.

## Data Availability

MIMIC IV database which was used in this study is openly available to credentialed users who sign 'Data Use Agreement' at PhysioNet website (20). The code is not publicly available due to privacy but a demo is available from the corresponding author on request.

## Acknowledgements

Not applicable.

## List of abbreviations

Abbreviation: **Full name**
DHP: Data Harmonisation Pipeline
EHR: Electronic Health Record
FHIR: Fast Healthcare Interoperability Resources
JSON: JavaScript Object Notation
MIMIC: Medical Information Mart for Intensive Care
RDF: Resource Description Format
XML: Extensible Markup Language

## Declarations

### Availability of data and materials

MIMIC IV database which was used in this study is openly available to credentialed users who sign “Data Use Agreement” at PhysioNet website (20). The code is not publicly available due to privacy but a demo is available from the corresponding author on request.

### Conflict of interest

The authors declare that they have no competing interests.

### Funding

This work was partially funded by the German Federal Ministry of Education and Research under Grant 16SV8559.

### Authors’ contributions

Study conception: EW, SN, MK, JR, AM

Data analysis: EW, MK

Figures: EW, SN, EM

Methods: EW, MK, AM, SN

Writing: EW, EM, JR, SAIK

Revising: BA, JB, PVB, JC, ARF, ASP, NS

## Competing Interests

The authors declare no competing interests.

## Notes

### Competing Interest Statement

The authors have declared no competing interest.

### Author Declarations

MIMIC IV database which was used in this study is openly available to credentialed users who sign 'Data Use Agreement' at PhysioNet website (20).

## References

1. Au-Yeung WTM, Sahani AK, Isselbacher EM, Armoundas AA. Reduction of false alarms in the intensive care unit using an optimized machine learning based approach. NPJ Digit Med [Internet]. 2019;2:86. Available from: https://europepmc.org/articles/PMC6728371

2. Desautels T, Calvert J, Hoffman J, Jay M, Kerem Y, Shieh L, et al. Prediction of Sepsis in the Intensive Care Unit With Minimal Electronic Health Record Data: A Machine Learning Approach. JMIR Medical Informatics. 2016 Jun;4.

3. Maier C, Kapsner L, Mate S, Prokosch HU, Kraus S. Patient Cohort Identification on Time Series Data Using the OMOP Common Data Model. Applied Clinical Informatics. 2021 Jun;12.

4. Ni Y, Wright J, Perentesis J, Lingren T, Deleger L, Kaiser M, et al. Increasing the efficiency of trial-patient matching: automated clinical trial eligibility Pre-screening for pediatric oncology patients. BMC Medical Informatics and Decision Making [Internet]. 2015;15(1):28. Available from: https://doi.org/10.1186/s12911-015-0149-3

5. de Mello BH, Rigo SJ, da Costa CA, da Rosa Righi R, Donida B, Bez MR, et al. Semantic interoperability in health records standards: a systematic literature review. Health and Technology [Internet]. 2022;12(2):255–72. Available from: https://doi.org/10.1007/s12553-022-00639-w

6. Kiourtis A, Mavrogiorgou A, Menychtas A, Maglogiannis I, Kyriazis D. Structurally Mapping Healthcare Data to HL7 FHIR through Ontology Alignment. Journal of Medical Systems. 2019 Jun;43.

7. Pagano P, Candela L, Castelli D. Data Interoperability. Data Sci J. 2013;12:GRDI19–25.

8. Rahm E, Bernstein P. A Survey of Approaches to Automatic Schema Matching. VLDB J. 2001 Jun;10:334–50.

9. Kolaitis P. Schema mappings, data exchange, and metadata management. In: Proceedings of the ACM SIGACT-SIGMOD-SIGART Symposium on Principles of Database Systems. 2005. p. 61–75.

10. HL7 FHIR. https://www.hl7.org/fhir/.

11. Vorisek C, Lehne M, Klopfenstein S, Bartschke A, Haese T, Thun S. Fast Healthcare Interoperability Resources (FHIR) for Interoperability in Health Research: A Systematic Review (Preprint). JMIR Medical Informatics. 2021 Jul;

12. Zong N, Wen A, Stone DJ, Sharma DK, Wang C, Yu Y, et al. Developing an FHIR-Based Computational Pipeline for Automatic Population of Case Report Forms for Colorectal Cancer Clinical Trials Using Electronic Health Records. JCO Clinical Cancer Informatics. 2020;4.

13. Hong N, Wen A, Shen F, Sohn S, Wang C, Liu H, et al. Developing a scalable FHIR-based clinical data normalization pipeline for standardizing and integrating unstructured and structured electronic health record data. JAMIA Open. 2019 Jun;2.

14. Martín Abadi, Ashish Agarwal, Paul Barham, Eugene Brevdo, Zhifeng Chen, Craig Citro, et al. TensorFlow: Large-Scale Machine Learning on Heterogeneous Systems [Internet]. 2015. Available from: https://www.tensorflow.org/

15. Paszke A, Gross S, Massa F, Lerer A, Bradbury J, Chanan G, et al. PyTorch: An Imperative Style, High-Performance Deep Learning Library. In: Advances in Neural Information Processing Systems 32 [Internet]. Curran Associates, Inc.; 2019. p. 8024–35. Available from: http://papers.neurips.cc/paper/9015-pytorch-an-imperative-style-high-performance-deep-learning-library.pdf

16. Liu D, Sahu R, Ignatov V, Gottlieb D, Mandl K. High Performance Computing on Flat FHIR Files Created with the New SMART/HL7 Bulk Data Access Standard. AMIA Annu Symp Proc. 2020 Mar 4;2019:592–6.

17. Oehm J, Storck M, Fechner M, Brix T, Yildirim K, Dugas M. FhirExtinguisher: A FHIR Resource Flattening Tool Using FHIRPath. In: Studies in health technology and informatics. 2021.

18. Mittelstadt B, Floridi L. The Ethics of Big Data: Current and Foreseeable Issues in Biomedical Contexts. Sci Eng Ethics. 2015 Jul;

19. Denney M, Long D, Armistead M, Anderson J, Conway B. Validating the Extract, Transform, Load Process Used to Populate a Large Clinical Research Database: International Journal of Medical Informatics. 2016 Jun;94.

20. Johnson A, Bulgarelli L, Pollard T, Horng S, Celi LA, Roger M. MIMIC-IV (version 2.0). PhysioNet. 2022.

21. PostgreSQL, PostgreSQL Global Development Group. https://www.postgresql.org. Accessed 15 June 2022.

22. Islam N. FHIR® Resources. https://github.com/nazrulworld/fhir.resources. Accessed 20 May 2022.

23. Konecný J, McMahan HB, Yu FX, Richtárik P, Suresh AT, Bacon D. Federated Learning: Strategies for Improving Communication Efficiency. ArXiv. 2016;abs/1610.05492.

24. Ulrich H, Behrend P, Wiedekopf J, Drenkhahn C, Kock-Schoppenhauer AK, Ingenerf J. Hands on the Medical Informatics Initiative Core Data Set — Lessons Learned from Converting the MIMIC-IV. In: Studies in health technology and informatics. 2021.

25. Bennett A, Wiedekopf J, Ulrich H, Johnson A. MIMIC-IV Clinical Database Demo on FHIR (version 2.0).. PhysioNet. 2022.

26. Shimabukuro DW, Barton CW, Feldman MD, Mataraso SJ, Das R. Effect of a machine learning-based severe sepsis prediction algorithm on patient survival and hospital length of stay: a randomised clinical trial. BMJ Open Respiratory Research [Internet]. 2017;4(1). Available from: https://bmjopenrespres.bmj.com/content/4/1/e000234

27. Sarmiento RF, Dernoncourt F. Improving Patient Cohort Identification Using Natural Language Processing. In: Data MITC, editor. Secondary Analysis of Electronic Health Records [Internet]. Cham: Springer International Publishing; 2016. p. 405–17. Available from: https://doi.org/10.1007/978-3-319-43742-2_28

28. Li J, Carayon P. Health Care 4.0: A vision for smart and connected health care. IISE Transactions on Healthcare Systems Engineering [Internet]. 2021;11(3):171–80. Available from: https://doi.org/10.1080/24725579.2021.1884627

29. McKinney W. Data Structures for Statistical Computing in Python. In 2010. p. 56–61.

