## Supplementary Information for "FHIR-DHP: A Standardized Clinical Data Harmonisation Pipeline for scalable AI application deployment"

**Supplementary Material**

Figure 1A. Flow chart showing an example diagnoses data saved in proposed custom JSON file being transformed to a “tensor” format. The diagnoses records saved in custom JSON(a) are first flattened to a tabular form (b), and then transformed into “tensor” (c).

In this section we would like to demonstrate how the proposed output custom JSON format can be easily transformed into an input data for common AI frameworks. To achieve a “tensor” shape we applied a two-step transformation. As shown in **Figure 1A.a**, an example diagnoses records are exported a custom AI-friendly format. In the first transformation step (see **Figure 1A.b**) the JSON format is flattened into a tabular form using *json_normalize* function from Pandas data preprocessing toolkit [1]. The second step, where the tabular data is transformed into a “tensor” form, is performed using *convert_to_tensor* function from the conventional AI framework Tensorflow [2].

**Supplementary References**

[1] McKinney W. Data Structures for Statistical Computing in Python. In 2010. p. 56–61.

[2] Martín Abadi, Ashish Agarwal, Paul Barham, Eugene Brevdo, Zhifeng Chen, Craig Citro, et al. TensorFlow: Large-Scale Machine Learning on Heterogeneous Systems [Internet]. 2015. Available from: https://www.tensorflow.org/
